# Diagnostic Performance of Pooled RT-PCR Testing for SARS-CoV-2 Detection

**DOI:** 10.1101/2021.02.17.21251961

**Authors:** Diadem Ricarte, Aubrey Gador, Leomill Mendiola, Ian Christian Gonzales

## Abstract

**Background:** With the high number of COVID-19 cases, a need to optimize testing strategy must be regarded to obtain timely diagnosis for early containment measures. With this, several studies have employed pooled RT-PCR testing for SARS-CoV-2 as this could potentially conserve laboratory resources while has the capacity to test several individuals. However, this was recommended to firstly validate the method as different laboratory reagents and equipment vary with its diagnostic performance.

**Objective:** The aim of this study was to determine the diagnostic performance of pooled SARS-CoV-2 nasopharyngeal/oropharyngeal swabbed samples using RT-PCR technique.

**Methods:** A records review of two-staged pooled RT-PCR testing data from August 10, 26, 30 and September 5, 2020 was utilized from Northern Mindanao Medical Center COVID-19 Satellite Laboratory (formerly CHDNM TB Regional Center). For the first stage, using known samples, a total of 30 pools were made for each of the pooling size, 5- and 10-pooled, on both pooling phase, pre- and post-RNA extraction. One positive individual was used to represent each of the Cycle threshold values given (<24, 25-28, 29-32, 33-36, and 37-40) while the rest of the samples were negative. For the second stage, 54 pools of five from 270 random unknown samples were used to validate the results. Target gene performance of N gene and RdRp was also determined.

**Key Results:** Results show that 5-pooled sample has higher sensitivity (SN), specificity (SP), positive predictive value (PPV), and negative predictive value (NPV) of 100% (95% confidence interval (CI) 88.97-100), 66.95% (95% CI, 60.75-72.6), 28.18% (95% CI, 20.62-37.22), and 100% (95% CI, 97.66-100) compared to 10-pooled sample that has 87.1% (95% CI, 71.15-94.87), 56.9% (50.57-63.02), 20.77% (95% CI, 14.68-28.53) and 97.14% (95% CI, 92.88-98.88). Further, these Ct values were only from the N gene, emphasizing its higher diagnostic performance as well to detect SARS-CoV-2 compared to RdRp as only a few samples were detected, thus, no analysis was made.

**Conclusion:** This study found out that 5-pooled sample has better diagnostic performance compared to 10-pooled samples. Specifically, all positive individual samples were detected in 5-pooled samples in pre-RNA extraction phase which these results are evident and consistent on both known and unknown samples. N gene was found out to detect more SARS-CoV-2 samples compared to RdRp.

## BACKGROUND

Coronavirus disease 2019, or known as COVID-19, was caused by a novel coronavirus, SARS-CoV-2, originating from the province of Hubei, Wuhan, China (Song, 2020). On January 30, 2020, the World Health Organization (2020) addressed this disease as a public health emergency of international concern as 32,730,945 confirmed COVID-19 cases including 958,703 deaths were tallied as of September 27, 2020. In the case of the Philippines, it has a total of 307,221 cases with 3,077 of these were detected in Northern Mindanao (Department of Health [DOH], 2020).

Detecting SARS-CoV-2 as early as possible displays a fundamental factor in containing its spread. However, with the unprecedented rise of COVID-19 cases, it caused a sudden huge strain on lives and economies as well as on the part of the healthcare system where a demand to upscale testing capacity is needed. While Philippines has now a total of 100 licensed reverse-transcriptase polymerase chain reaction (RT-PCR) laboratories as of August 7, 2020 with an average daily testing capacity of 27,800 in the last seven days (DOH, 2020), it still faces a challenge on the constant supply of laboratory resources as procurement has been difficult due to international competition (Galvez, 2020). In addition, though the use of RT-PCR diagnostic test is highly recommended to detect SARS-CoV-2, the shortage of supplies hampers the need to continually operate and overwhelm finances as well (Levenson, 2020; Owens-Liston, 2020; Scott, 2020). As many are experiencing this crisis, an alternative way of detecting the virus through optimizing strategies to conserve diagnostic kits while still having the capacity to increase testing must be regarded.

The European Center for Disease Control and Prevention (2020) has recommended the use of a pooled testing method where it aids in monitoring the activity of a disease whilst providing a rational use of diagnostic kits especially in low-resource settings (Majid et al., 2020). Also, this strategy can also expand the capacity of the laboratory to conduct surveillance testing (US Centre for Disease Prevention and Control [US CDC], 2020; Rohde, 2020).

Several studies have already employed the use of a pooled testing method to detect SARS-CoV-2 samples (Hogan et al., 2020; Lohse et al., 2020; Bi et al., 2020). However, this has also been recommended to firstly validate as different laboratory reagents and equipment vary with its diagnostic performance (Abdalhamid, 2020), specifically in terms of its sensitivity, specificity, positive predictive value, and negative predictive value. In the context of Northern Mindanao, Philippines, this method was explored in this study, specifically addressed the question, “What is the diagnostic performance of pooled SARS-CoV-2 nasopharyngeal/oropharyngeal swabbed samples using RT-PCR technique?”

With the rising cases of COVID-19 that resulted in shortage of laboratory supplies, this study addresses the need to increase testing capacity in a cost-effective manner as supply problems are globally experienced due to the pandemic. Since this method is not feasible for SARS-CoV-2 standard testing, this approach can be a potential tool to use for epidemiological reasons such as determining the proportion of positive cases within a certain area in the region as local transmission is also occurring. Most especially, Philippines, despite its current capacity, is not testing enough. With this pooled testing method, if proven to be effective, will allow more individuals to be tested and their COVID-19 viral status known, thus, it could potentially improve disease prevention and control strategies of the government such as aid for early detection of positive cases while assisting policymakers’ decisions for their implementation of containment measures. While Northern Mindanao is experiencing a delay in supply logistics that resulted in a backlog of samples, hence, this pooling method can possibly conserve the resources and continue testing individuals.

## METHODOLOGY

A retrospective research design was employed to obtain secondary data on SARS-CoV-2 swabbed samples that were collected from nasopharyngeal and oropharyngeal sites of individuals ranging from asymptomatic, mild with no comorbidities, mild with comorbidities, severe and critical cases in Northern Mindanao and that were sent by the trained Medical Technologists to the Northern Mindanao Medical Center COVID-19 Satellite Laboratory (formerly CHDNM TB Regional Center) last August 10, 26, 30, 2020 and September 5, 2020.

### Inclusion criteria

- Known and unknown SARS-CoV-2 sample results within the valid control were included for data collection where a positive control had a Ct value of ≤22 for RdRp and N gene while a Ct value of ≤21 for RNAse P (Internal control) and no detection cycles of the genes mentioned for a negative control as also based on the Instruction for Use (IFU) of GeneFinder™ COVID-19 Plus RealAmp Kit. For further validation, the pathologist, two Medical Technologists as well as the Head of the Infectious Disease Cluster verified the control results.
- Individual and pooled samples that were received and tested within 24 hours of receipt to Northern Mindanao Medical Center COVID-19 Satellite Laboratory (formerly CHDNM TB Regional Center) were only included for data collection.

### Exclusion criteria

- GeneFinder™ COVID-19 Plus RealAmp Kit uses three target genes: N gene, E gene and RdRp and RNAse P as the internal control gene. However, E gene is only interpreted as PRESUMPTIVE POSITIVE if it is detected, thus, this gene was not included for the data collection.

### Sampling Method

This study had two stages. First, the pooling data on known (determined) samples was used to identify what ideal sample size and pooling phase could detect the positive samples in each of the Ct value ranges given. Its target genes and diagnostic performance were analyzed. Second, the pooling data on arbitrarily chosen unknown (clinical) samples was then used to validate the ideal results found on the pooled known samples in which its target genes and diagnostic performance were analyzed.

The methodology on how the pooled samples were done prior and after RNA extraction were based on the published literature (Abdalhamid et al., 2020; Gupta et al., 2020; Perchetti et al., 2020; Yelin et al., 2020) having high diagnostic performance and reliable results, thus, its pooling procedures were synthesized and adopted in this study.

#### i. Sample preparation

- Combined nasopharyngeal and oropharyngeal swabs were collected by the trained DOH CHDNM Medical Technologists, transported at 2 to 8 degree Celsius in a 2 ml Sanli Virus specimen collection tube; Liuyang, Sanli Medical Technology Devt. Co., China (RITM, 2020), a valid and compatible for the SARS-CoV-2 detection kit used where it states that it is intended for use with RNA extracted from nasopharyngeal swabs, and oropharyngeal swabs. However, there was no ideal transport medium indicated in its Instruction for Use (GeneFinder™ COVID-19 Plus RealAmp Kit, OSANG Healthcare Co., Ltd, South Korea), thus, valid VTM is acceptable. The samples that were sent to the said laboratory were directly processed. While the results of the individual samples were yet to be determined for a turnaround time of 8 to 12 hours, VTM were stored at 4 degree Celsius. Once known, these were then pooled after. Hence, no freeze-thaw cycles were done.
- From the time the samples were received at the laboratory, these were then processed to avoid RNA degradation. Samples were labeled and assigned in a designated pool for each of the Ct value ranges given. After which, these were prepared in pool of 5 and pool of 10 and were pooled in two phases, pre-RNA extraction and post-RNA extraction in three replicates to ensure reproducibility of the results. For a pool of five, one positive and four negative samples were used in each of the Ct value ranges: <24, 25-28, 29-32, 33-36 and 37-40. For a pool of ten, one positive and nine negative samples were used in each of the Ct value ranges: <24, 25-28, 29-32, 33-36 and 37-40. (Supplementary Figure S1 and S2).
- For the first stage, pooling of known samples was used. Samples were pooled right after the results of the individual samples were determined or about 12 hours from the time individual samples were received at the laboratory and tested until these were then pooled. Previously tested samples were stored at 4 degree Celsius prior to pooling. For the second stage, pooling of unknown samples was used. Individual and pooled samples were tested in parallel.

#### ii. RNA Extraction

An automated extractor, Genolution Nextractor® NX-48S with its nucleic acid reagents, was used for the purification of RNA. The extraction underwent the following steps: lysis of the sample, DNA binding to the magnetic beads, washing and elution.

- Individual samples

A total volume of 200 microliter from each of the universal transport medium buffers was dispensed onto the pre-filled sample well of the automated Genolution Nextractor® NX-48S plate. After loading all the individual samples, these were placed into the auto extractor for RNA purification.

- Pooling of samples prior to RNA extraction

An initial volume of 100 microliter aliquot from each viral transport medium buffer was initially transferred onto a conical tube for the pooling of samples prior to dispensing onto the pre-filled sample well of the automated Genolution Nextractor NX-48S plate. A total volume aliquot of 500 microliter for the 5-pooled sample and 1,000 microliters for the 10-pooled samples. A final volume of 200 microliter, either of the 5- or 10-pooled samples, was dispensed into the pre-filled sample well of the automated Genolution Nextractor NX-48S plate. The samples were pooled in three replicates. Eluted samples were then transferred to a new tube for RT-qPCR test.

- Pooling of samples after RNA extraction

Eluted RNA samples were firstly pooled prior to mixing with the master mix reagent onto the 96-well plate. An aliquot volume of 10 microliter each eluted RNA sample was dispensed initially onto the tube for pooling. A total volume of 50 microliter for the 5-pooled samples and 100 microliters for the 10-pooled samples. A final volume of 5 microliter, either of the 5- or 10-pooled samples, was dispensed onto the 96-well plate for RT-PCR testing.

#### iii. RT-PCR Testing

After the RNA extraction, RT-PCR test was performed to detect the presence of SARS-CoV-2 RNA using GeneFinder™ COVID-19 Plus RealAmp Kit in Applied Biosystems® 7500 Real-Time PCR Instrument (ABI 7500) machine (Thermo Fisher Scientific Inc.) based on the Instruction for Use (IFU) provided.

A 15 microliter of GeneFinder™ master mix reagent was dispensed onto the 96-well plate. After which, a 5 microliter of eluted samples, either of the individual, 5- or 10-pooled samples, were dispensed. A 96-well plate containing mastermix reagent and eluted RNA sample was placed onto the RT-PCR machine to detect 3 SARS-CoV-2 target genes: N, E, and RdRP. For quality control, there were three controls provided on the Instruction for Use (IFU) of GeneFinder™ COVID-19 Plus RealAmp Kit. For internal control, RNAse P gene with Ct value of ≤22 was used while for positive external control, N gene and RdRp with Ct value of ≤22. No detection cycles of the genes mentioned was set for a negative control.

Reactions were heated to 50° for 20 minutes for reverse transcription, denatured at 95° for 15 minutes and then 45 cycles were carried for amplification set at 95° for 15 seconds and 58° for 1 minute. Fluorescence was measured using four channels: FAM (RdRp gene), JOE (N gene), TEXAS RED (E gene), and CY5 (RNAse P). After which, the data on the individual and pooled SARS-CoV-2 NP/OP swabbed samples were validated and will be obtained for analysis.

## RESULTS AND DISCUSSION

### Study Samples

#### Pooling of previously known samples

A total of 450 known samples were utilized for this stage. A positive sample was used to represent each of the cycle threshold values given. There were 150 known samples used for pooling of five while 300 samples were used for pooling of 10 that consist of one positive sample and four negative samples in each of the five Ct value ranges and were pooled in two phases, prior to RNA extraction or after RNA extraction, in three replicates.

##### **i**. Pooling of 5-pooled samples

Table 1 shows that all five positive individual samples were detected when pooled in pre-RNA extraction phase only. On the other hand, a positive sample with an N gene Ct value of 37.66 (Ct value range 37-40) was negative when pooled in post-RNA extraction phase (Ct Mean: 41.85; SD: 7.94). Meanwhile, no analysis for RdRp Ct values was made as it was undetected for most of the samples (Table 2).

**Table 1.** Cycle threshold values of N gene in previously known individual and 5-pooled samples in the given RNA extraction phase.

| Ct value range | Individual Samples<sup>a</sup> | Pooled Samples |  |
| --- | --- | --- | --- |
|  |  | Pre RNA extraction | Post RNA extraction |
|  |  | Mean (SD)* | Mean (SD)* |
| <24 | 17.6 | 19.14 (0.87) | 21.51 (0.71) |
| 25-28 | 25 | 26.08 (0.62) | 28.00 (0.06) |
| 29-32 | 31.26 | 31.01 (0.53) | 34.12 (2.52) |
| 33-36 | 33.23 | 32.29 (0.03) | 33.98 (1.80) |
| 37-40 | 37.66 | 34.82 (2.15) | 41.85 (7.94)<sup>b</sup> |

<sup>a</sup>Individual samples refer to the Ct values of known positive samples only
<sup>b</sup>SARS-CoV-2 is not detected
SD: Standard deviation
\*done in triplicates

**Table 2.** Frequency of RdRp gene in 5-pooled samples in the given RNA extraction phase.

| Pre RNA-Extraction |  | Post-RNA Extraction |  |
| --- | --- | --- | --- |
| Detected | Undetected | Detected | Undetected |
| 3 | 12 | 2 | 13 |

##### **ii**. Pooling of 10-pooled samples

Results show that one positive individual sample with a Ct value of 37.66 (Ct value range: 37-40) cannot be detected when pooled on both RNA extraction phases (Table 3). Meanwhile, no analysis for RdRp Ct values was made as it was undetected for most of the samples (Table 4).

**Table 3.** Cycle threshold values of N gene in previously known individual and 10-pooled samples in the given RNA extraction phase.

| Ct value range | Individual Samples<sup>a</sup> | Pooled Samples |  |
| --- | --- | --- | --- |
|  |  | Pre RNA extraction | Post RNA extraction |
|  |  | Mean (SD)* | Mean (SD)* |
| <24 | 17.6 | 20.48 (0.36) | 22.24 (0.06) |
| 25-28 | 25 | 27.16 (1.15) | 29.40 (0.16) |
| 29-32 | 31.26 | 32.76 (1.80) | 33.38 (0.88) |
| 33-36 | 33.23 | 35.02 (1.21) | 34.62 (0.45) |
| 37-40 | 37.66 | - | - |

<sup>a</sup>Individual samples refer to the Ct values of known positive samples only
SD: Standard deviation
\*done in triplicates

**Table 4.** Frequency of RdRp gene in 10-pooled samples in the given RNA extraction phase.

| Pre RNA-Extraction |  | Post-RNA Extraction |  |
| --- | --- | --- | --- |
| Detected | Undetected | Detected | Undetected |
| 1 | 14 | 1 | 14 |

#### Validation of pool testing using random unknown samples

As all known individual positive samples can only be detected when these are pooled in five in pre-RNA extraction phase, the results were validated with 270 unknown samples. These were sorted in 54 pools. Results show that all 31 samples that tested positive on individual testing, which got sorted into 22 pools, were also detected in pooled sample testing. These positive pools were composed of one positive sample (14/22), two positive samples (7/22), and three positive samples (1/22) (Supplementary Table S1).

## Main Results of the Study

### Difference between pooling sizes in two pooling phases of the given Cycle threshold value ranges

To compare the difference between 5-pooled and 10-pooled known nasopharyngeal/oropharyngeal swabbed samples that is pooled in each RNA extraction phase in detecting SARS-CoV-2 in Ct value ranges of <24, 25-28, 29-32, 33-36, 37-40, Open Epi was used to analyze the set of data.

Table 5 shows that there is no significant difference between 5- and 10-pooled samples in Ct value range of 29-32 (95 CI: −0.68, 2.16) and 33-36 (95 CI: −1.65, 0.37) in post-RNA extraction phase. Further, a known positive sample in Ct value range of 37-40 is negative in 5-pooled sample of post-RNA extraction phase and is undetected in 10-pooled samples for both RNA extraction phases.

**Table 5:** N gene mean Ct values of pooled samples in pre- and post-extraction phase.

| Ct value range | Pre-RNA Extraction |  |  | Post-RNA Extraction |  |  |
| --- | --- | --- | --- | --- | --- | --- |
|  | 5-pooled | 10-pooled | 95 CI of difference | 5-pooled | 10-pooled | 95 CI of difference |
| <24 | 19.14 | 20.48 | (-1.84, -0.84) | 21.51 | 22.24 | (-1.12, -0.34) |
| 25-28 | 26.08 | 27.16 | (-1.61, -0.55) | 28 | 29.4 | (-1.47, -1.33) |
| 29-32 | 31.01 | 32.76 | (-2.47, -1.03) | 34.12 | 33.38 | (-0.68, 2.16) |
| 33-36 | 32.29 | 35.02 | (-3.18, -2.28) | 33.98 | 34.62 | (-1.65, 0.37) |
| 37-40 | 34.82 | - | - | 41.85 | - | - |

### Performance of the target genes in two pooling sizes in pre-RNA extraction phase

To compare the diagnostic performance between the target genes, N gene and RdRp, in detecting unknown 5-pooled and 10-pooled SARS-CoV-2 nasopharyngeal/oropharyngeal swabbed samples that is pooled in pre-RNA extraction phase, Microsoft Excel and Open Epi was used to analyze the set of data.

Using 2×2 table analysis (Table 6, 7, 9, and 10), results show that N gene of 5-pooled sample has higher diagnostic performance in terms of its sensitivity, positive predictive value, and negative predictive value. On the other hand, RdRp of 5-pooled sample is more specific. However, there is no significant difference between the target genes, N gene and RdRp, for both pooling sizes, 5-pooled and 10-pooled samples, in detecting SARS-CoV-2 nasopharyngeal/oropharyngeal swabbed samples (Table 8 and 11).

**Table 6.** 2×2 table analysis of individual samples and 5-pooled N gene.

|  |  | Individual samples |  |  |
| --- | --- | --- | --- | --- |
| 5-pooled N gene |  | (+) | (-) | Total |
|  | (+) | 30 | 80 | 110 |
|  | (-) | 1 | 159 | 160 |
|  |  | 31 | 239 | 270 |

**Table 7.**
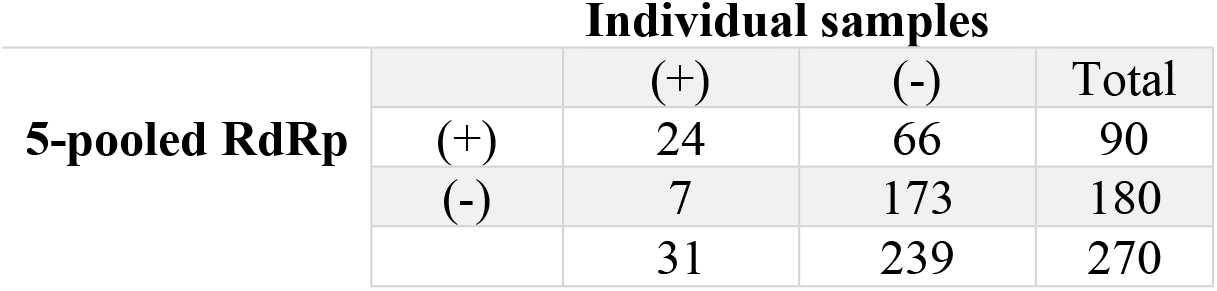
2×2 table analysis of individual samples and 5-pooled RdRp.

**Table 8.**
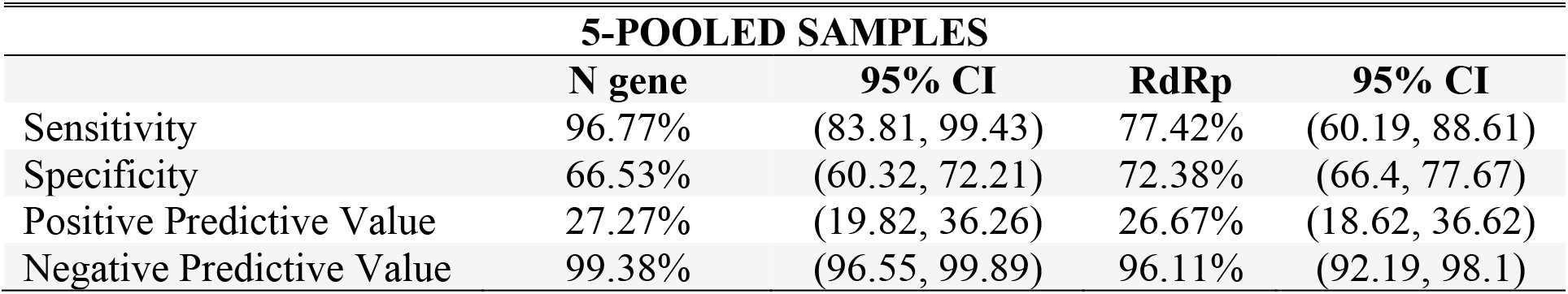
Diagnostic performance of target genes of 5-pooled samples.

**Table 9.** 2×2 table analysis of individual samples and 10-pooled N gene.

|  |  | Individual samples |  |  |
| --- | --- | --- | --- | --- |
| 10-pooled N gene |  | (+) | (-) | Total |
|  | (+) | 25 | 75 | 100 |
|  | (-) | 6 | 164 | 170 |
|  |  | 31 | 239 | 270 |

**Table 10.** 2×2 table analysis of individual samples and 10-pooled RdRp.

|  |  | Individual samples |  |  |
| --- | --- | --- | --- | --- |
| 10-pooled RdRp |  | (+) | (-) | Total |
|  | (+) | 27 | 103 | 130 |
|  | (-) | 4 | 136 | 140 |
|  |  | 31 | 239 | 270 |

**Table 11.** Diagnostic performance of target genes of 10-pooled samples.

| **10-POOLED SAMPLES** |  |  |  |  |
| --- | --- | --- | --- | --- |
|  | **N gene** | **95% CI** | **RdRp** | **95% CI** |
| Sensitivity | 87.1% | (71.15, 94.87) | 80.65% | (63.72, 90.81) |
| Specificity | 56.9% | (50.57, 63.02) | 68.62% | (62.48, 74.17) |
| Positive Predictive Value | 20.77% | (14.68, 28.53) | 25.00% | (17.55, 34.3) |
| Negative Predictive Value | 97.14% | (92.88, 98.88) | 96.47% | (92.51, 98.37) |

### Performance of the two pooling sizes in pre-RNA extraction phase

To compare the diagnostic performance of the pooling method of unknown 5-pooled and 10-pooled SARS-CoV-2 nasopharyngeal/oropharyngeal swabbed samples that is pooled in pre-RNA extraction phase, Microsoft Excel and Open Epi was used to analyze the set of data.

Using 2×2 table analysis (Table 12 and 13), Table 14 shows that 5-pooled samples have higher diagnostic performance compared to 10-pooled samples. However, there is no significant difference between the two pooling sizes.

**Table 12.** 2×2 table analysis of individual and 5-pooled samples.

| **Individual samples** |  |  |  |  |
| --- | --- | --- | --- | --- |
| **10-pooled** |  | (+) | (-) | Total |
|  | (+) | 31 | 79 | 110 |
|  | (-) | 0 | 160 | 160 |
|  |  | 31 | 239 | 270 |

**Table 13.** 2×2 table analysis of individual and 10-pooled samples.

| **Individual samples** |  |  |  |  |
| --- | --- | --- | --- | --- |
| **10-pooled** |  | (+) | (-) | Total |
|  | (+) | 27 | 103 | 130 |
|  | (-) | 4 | 136 | 140 |
|  |  | 31 | 239 | 270 |

**Table 14.** Diagnostic performance of 5- and 10-pooled samples.

| **POOLED SAMPLES** |  |  |  |  |
| --- | --- | --- | --- | --- |
|  | **5-pooled** | **95% CI** | **10-pooled** | **95% CI** |
| Sensitivity | 100% | (88.97, 100) | 87.1% | (71.15, 94.87) |
| Specificity | 66.95% | (60.75, 72.6) | 56.9% | (50.57, 63.02) |
| Positive Predictive Value | 28.18% | (20.62, 37.22) | 20.77% | (14.68, 28.53) |
| Negative Predictive Value | 100% | (97.66, 100) | 97.14% | (92.88, 98.88) |

## Discussion

This study found out that 5-pooled sample in pre-RNA extraction phase has higher diagnostic performance as results are evident and consistent on both known and unknown samples. To further elaborate, it has a sensitivity (SN), specificity (SP), positive predictive value (PPV), and negative predictive value (NPV) of 100% (95% confidence interval (CI) 88.97-100), 66.95% (95% CI, 60.75-72.6), 28.18% (95% CI, 20.62-37.22), and 100% (95% CI, 97.66-100) compared to 10-pooled sample that has 87.1% (95% CI, 71.15-94.87), 56.9% (50.57-63.02), 20.77% (95% CI, 14.68-28.53) and 97.14% (95% CI, 92.88-98.88), respectively (Table 8 and 11). However, the diagnostic performance of SARS-CoV-2 pooling method by Abdalhamid et. al. (2020) which has SN of 95% and SP of 100% using five samples and de Salazar et. al. (2020) which has SN, SP, PPV and NPV of 97.10% (95% CI, 94.11-98.82), 100%, 100% and 99.79% (95% CI, 99.56-99.90) on pooling of ten samples were higher than the results of this study. This may probably be due to higher sensitivity of the viral detection kit used as it potentially contribute on the diagnostic performance of the pooling method.

In this study, the higher diagnostic performance of 5-pooled sample is expected as these were diluted less than 10-pooled which tends to increase the cycle threshold value and lowers the viral load. This is also apparent when pooling was firstly done on known samples as only 5-pooled in pre-RNA extraction phase could be able to detect all known individual positive samples, specifically the N gene, on all the cycle threshold value ranges given (Table 1 & 5). According to Abdalhamid et. al. (2020), pooling of five samples is also efficient as it could yield a 69% increase in testing capability, highlighting the importance of the method to address the shortage of supplies. These findings are also similar with the study of Chhikara et. al. (2020) and Alcoba-Florez et. al. (2020) where they concluded that pooling of 5 samples is sufficient to detect the presence of SARS-CoV-2, especially when pooled in pre-RNA extraction phase (Alcoba-Florez et al., 2020; Volpato et al., 2020). However, this is in contrary to Yelin et. al. (2020) that positive samples can be detected in pool of up to 32 samples as this study shows that only five samples can be pooled efficiently. The prevalence rate of a certain area must also be considered as this contributes to the practicality and its cost-effectiveness (Dorfman, 1943) as a range of 3 to 10 pool sizes in a two-stage pooling algorithm can be used if there is a prevalence rate of 5% (Abdalhamid et al., 2020). Further, pooling of samples prior to RNA significantly portrays that a high number of diagnostic resources may be conserved compared when samples are pooled after RNA extraction as it causes delay of individual sample detection, as positive pools need to be deconvoluted and is also prone to degradation of protein level. There were only limited studies who conducted pooled testing on both pre- and post-RNA extraction phases (Chhikara et. al., 2020; Volpato et. al., 2020; Mulu et. al., 2020) thus, this study has added knowledge on this concept.

As shown in this study, only N gene was detected on all known individual positive samples when pooled in five in pre-RNA extraction phase. When validated and analyzed on unknown samples, it was found out that this gene is more sensitive and has higher positive predictive value and negative predictive value, however, RdRp is more specific. To the best of our knowledge, target gene performance was only evaluated on individual samples. According to Benrahma et. al. (2020), N gene is persistently detected in samples with Ct value of ≤ 40 in RT-PCR having similar viral detection kit used in this study. This may explain due to its abundant production within infected cells having multiple functions such as replication, transcription, translation and specifically binding to viral RNA to form the ribonucleocapsid (McBride, Van Zyl and Fielding, 2014). A comparative study of Vogels et. al. (2020) and Nalla et. al. (2020) also showed that N gene is found to be more sensitive than RdRp when tested on individual nasopharyngeal and oropharyngeal samples, and noticeably more when it is pooled as illustrated in this study. In addition, this is also emphasized when an evaluation of SARS-CoV-2 detection kit was done by Zhou et. al. (2020) and recommended by the study of Chu et. al. (2020) as a screening assay as N gene is ten times more sensitive than Orf1b in detecting positive samples. On the other hand, as RdRp is a viral enzyme with no host cell homolog that functions for viral RNA replication, this might explain why it has higher diagnostic performance (Zhu, et al., 2020).

The scope of this study is the diagnostic performance of the reagents and equipment used. Further, its limitation includes no analysis on the pooling of SARS-CoV-2 swabbed samples of the symptomatic and asymptomatic individuals as only de-identified samples were utilized. Overall, this study serves as a proof of concept of the pooling method, illustrating the results of the different pooling sizes on both RNA extraction phases, of SARS-CoV-2 using RT-PCR technique. In conclusion, this study found out that 5-pooled sample has better diagnostic performance compared to 10-pooled samples as, specifically, all positive individual samples were only detected in 5-pooled samples in pre-RNA extraction phase which these results are evident and consistent on both known and unknown samples. Further, N gene was found out to detect more SARS-CoV-2 samples compared to RdRp.

Based on the results of this study, the following are hereby recommended for future research and development work:

For research institutes

- Given the scope of this study, a validation on pooling method using other diagnostic reagents and equipment must also be done as its performance may vary and may not be similar with the results shown in this study.
- To address the limitation of this study, consider symptom profile and its duration when pooling of SARS-CoV-2 swabbed samples.
- If incidence or surveillance of COVID-19 is aimed for the study, a pooled testing method is suggested to lessen the workload and resources used while at the same time having a reliable result compared to using rapid diagnostic kits. The results may also be a significant basis for policy-making in the respective areas.

For policymakers:

- Based on the results of the study, pooling of five samples prior to RNA extraction must be used as it could possibly detect much positive samples even for a high cycle threshold value or low viral load compared to pooling of 10 samples that has a higher chance of missing out possible positive samples. Moreover, this could be a potential tool to hasten the process of tracking and isolating possible COVID-19 individuals.
- For pooling of SARS-CoV-2, the following reagents and equipment are hereby suggested as these shows satisfactory performance: Genolution Nextractor® NX-48S for RNA extraction, GeneFinder™ COVID-19 Plus RealAmp Kit for SARS-CoV-2 viral detection and Applied Biosystems® 7500 Real-Time PCR Instrument (ABI 7500; Thermo Fisher Scientific Inc.) for PCR machine.

## Data Availability

The data that support the findings of this study are available from the corresponding author upon reasonable request.

## APPENDICES

### I. Pooling Procedure

**Supplementary Figure S1.**
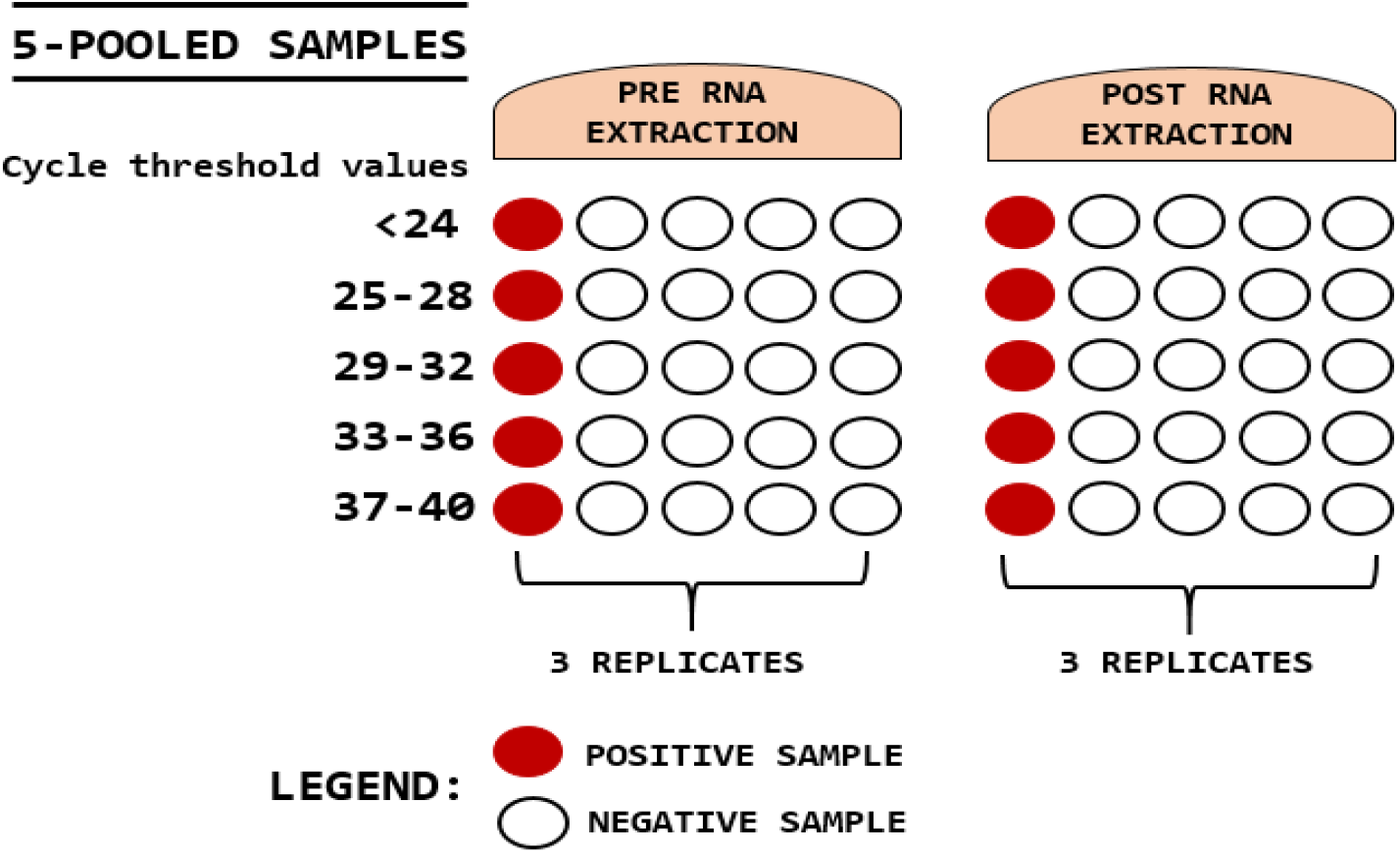
Illustration of 5-pooled samples with one positive sample (red circle) and four negative samples (white circle) in each of the Ct value ranges given and pooled in two phases: pre-RNA extraction and post RNA extraction. These were done in three replicates.

**Supplementary Figure S2.**
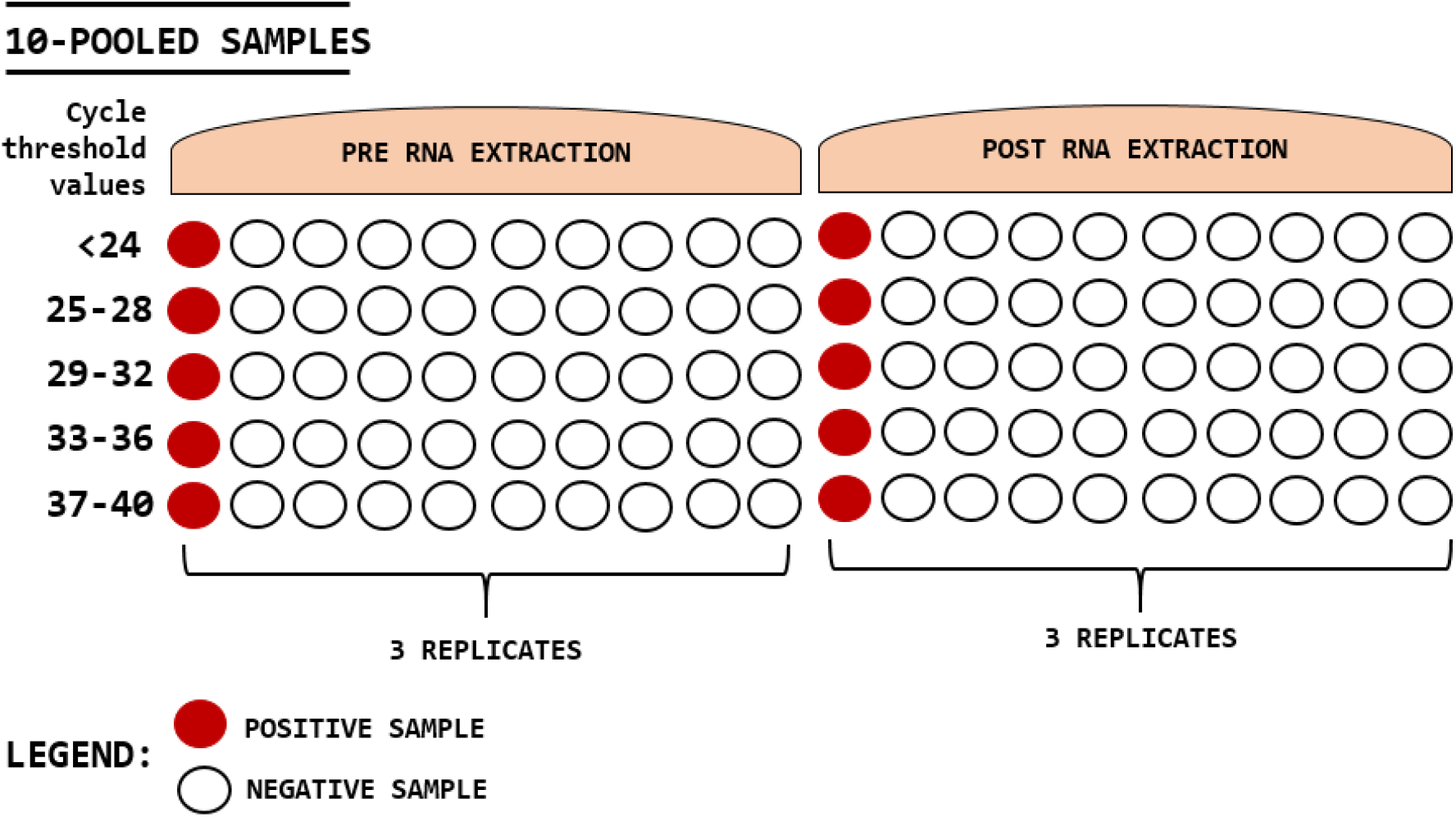
Illustration of 10-pooled samples with one positive sample (red circle) and nine negative samples (white circle) in each of the Ct value ranges given and pooled in two phases: pre-RNA extraction and post RNA extraction. These were done in three replicates.

### II. Raw data and statistical formula and analysis

**Supplementary Table S1.** Cycle threshold values of individual positive samples in 5-pooled pre-RNA extraction phase.

| Positive Sample No. | Pool ID | Individual samples |  | 5-pooled samples |  |
| --- | --- | --- | --- | --- | --- |
|  |  | N gene | RdRp | N gene | RdRp |
| 1 | A5 | 19.06 | 19.85 | 21.21 | 21.84 |
| 2 |  | 17.12 | 17.31 | 21.21 | 21.84 |
| 3 | E5 | 37.33 | 40.8 | 22.34 | 22.16 |
| 4 |  | 19.5 | 19.51 | 22.34 | 22.16 |
| 5 | G5 | 35.87 | 40.95 | 36.04 | 40.21 |
| 6 | I5 | 25.51 | 25.75 | 23.68 | 23.61 |
| 7 |  | 25.29 | 26.57 | 23.68 | 23.61 |
| 8 | J5 | 27.24 | 28.88 | 23.28 | 23.52 |
| 9 | K5 | 32.84 | 35.8 | 37.14 | 39.01 |
| 10 | P5 | 24.34 | 29.07 | 22.51 | 23.12 |
| 11 | S5 | 33.39 | 36.15 | 31.88 | 32.78 |
| 12 | T5 | 23.87 | 24.39 | 26.72 | 27.38 |
| 13 |  | 27.06 | 29.03 | 26.72 | 27.38 |
| 14 | U5 | 27.42 | 24.35 | 26.06 | 25.32 |
| 15 | V5 | 23.68 | 24.43 | 28.25 | 28.94 |
| 16 | W5 | 30.22 | 30.49 | 24.23 | 23.92 |
| 17 |  | 21.01 | 20.58 | 24.23 | 23.92 |
| 18 | X5 | 21.97 | 21.3 | 24.36 | 25.61 |
| 19 | Y5 | 34.67 | 35.2 | 37.62 | UND |
| 20 |  | 35.82 | 34.76 | 37.62 | UND |
| 21 |  | 33 | 34.13 | 37.62 | UND |
| 22 | Z5 | 28.97 | 29.66 | 31.69 | 32.45 |
| 23 | B51 | 31.34 | 31.77 | 35.44 | 36.94 |
| 24 | C51 | 37.11 | UND | 37.96 | UND |
| 25 |  | 35.91 | 40.61 | 37.96 | UND |
| 26 | D51 | 31.1 | 32.4 | 33.2 | 33.72 |
| 27 |  | 32.44 | 32.73 | 33.2 | 33.72 |
| 28 | G51 | 31.71 | 32.76 | 35.42 | 41.5 |
| 29 | N51 | 33.73 | 34.01 | 36.05 | 37.61 |
| 30 | X51 | 38.84 | 37.26 | 35.89 | 38.97 |
| 31 | Y51 | UND | 37.65 | 37 | 38.7 |

UND: Viral gene is undetected

**Supplementary Table S2.** Comparison of mean Ct value of 5- and 10-pooled samples in pre-extraction phase with a Ct value of <24.

| TWO-SAMPLE INDEPENDENT *t* TEST |  |  |  |  |  |  |
| --- | --- | --- | --- | --- | --- | --- |
| Two-sided confidence interval |  | 95% |  |  |  |  |
|  | Sample size | Mean | Std. Dev. |  |  |  |
| 5-pooled | 15 | 19.14 | 0.87 |  |  |  |
| 10-pooled | 30 | 20.48 | 0.36 |  |  |  |
| **Result** | ***t* statistics** | ***df*** | **p-value<sup>1</sup>** | **Mean Difference** | **Lower Limit** | **Upper Limit** |
| Equal variance | -7.33394 | 43 | <0.0000001 | -1.34 | -1.70848 | -0.971524 |
| Unequal variance | -5.72524 | 16 | 0.00003129 | -1.34 | -1.83616 | -0.843837 |
|  |  | ***F* statistics** | ***df*(numerator,denominator)** | **p-value<sup>1</sup>** |  |  |
| Test for equality of variance<sup>2</sup> |  | 5.84028 | 14,29 | 0.00006244 |  |  |

<sup>1</sup> p-value (two-tailed)

**Supplementary Table S3.**
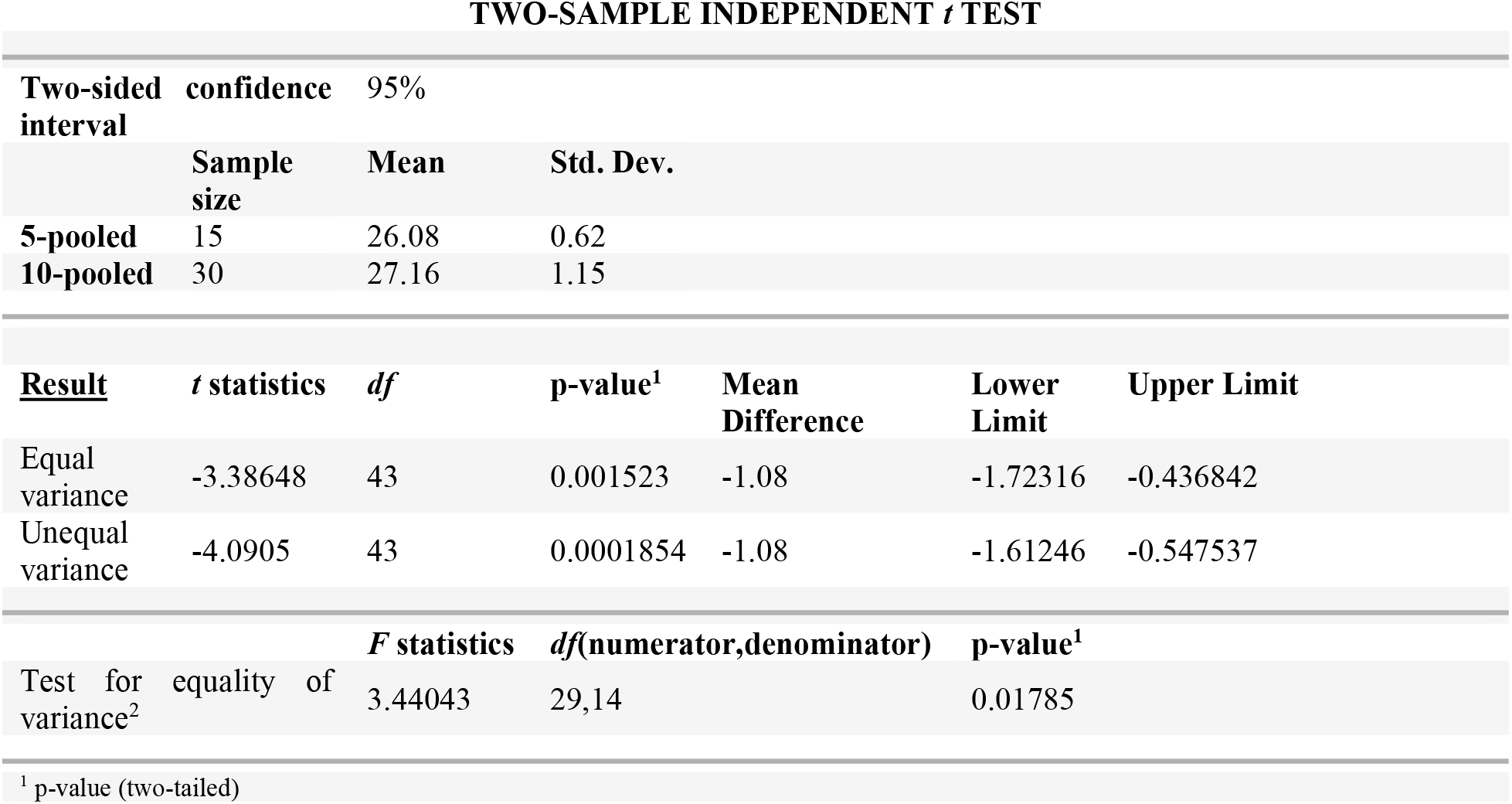
Comparison of mean Ct value of 5- and 10-pooled samples in pre-extraction phase with a Ct value of 25-28.

**Supplementary Table S4.**
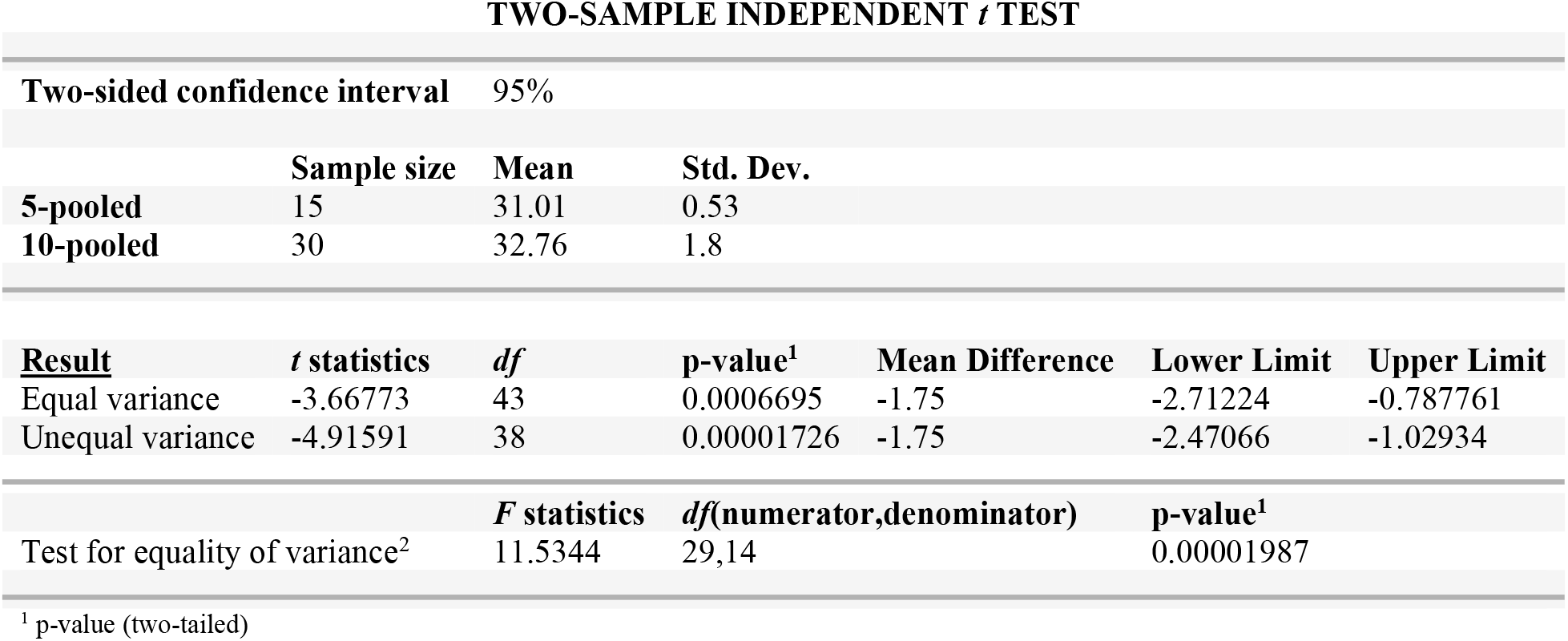
Comparison of mean Ct value of 5- and 10-pooled samples in pre-extraction phase with a Ct value of 29-32.

**Supplementary Table S5.**
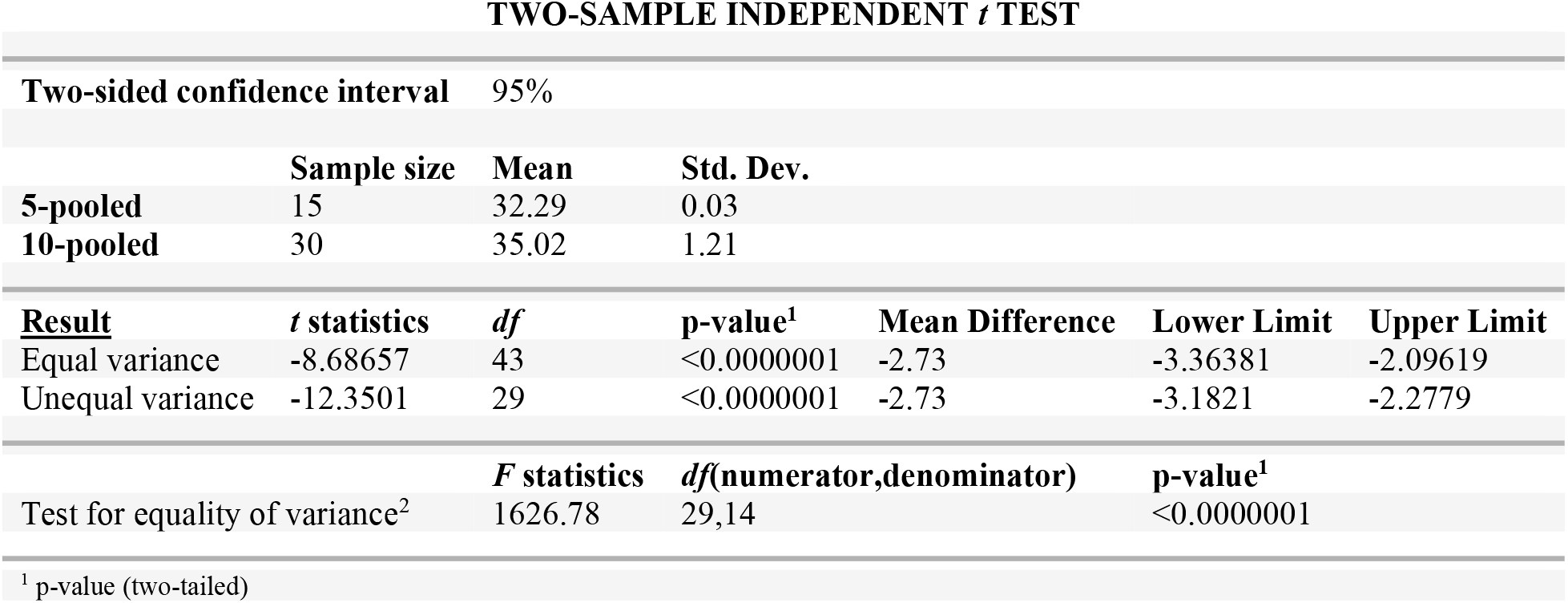
Comparison of mean Ct value of 5- and 10-pooled samples in pre-extraction phase with a Ct value of 33-36.

| TWO-SAMPLE INDEPENDENT *t* TEST |  |  |  |  |  |  |
| --- | --- | --- | --- | --- | --- | --- |
| Two-sided confidence interval |  | 95% |  |  |  |  |
|  | Sample size | Mean | Std. Dev. |  |  |  |
| 5-pooled | 15 | 32.29 | 0.03 |  |  |  |
| 10-pooled | 30 | 35.02 | 1.21 |  |  |  |
| **Result** | ***t* statistics** | ***df*** | **p-value<sup>1</sup>** | **Mean Difference** | **Lower Limit** | **Upper Limit** |
| Equal variance | -8.68657 | 43 | <0.0000001 | -2.73 | -3.36381 | -2.09619 |
| Unequal variance | -12.3501 | 29 | <0.0000001 | -2.73 | -3.1821 | -2.2779 |
|  | ***F* statistics** | ***df*(numerator,denominator)** |  | **p-value<sup>1</sup>** |  |  |
| Test for equality of variance<sup>2</sup> | 1626.78 | 29,14 |  | <0.0000001 |  |  |

<sup>1</sup> p-value (two-tailed)

**Supplementary Table S6.**
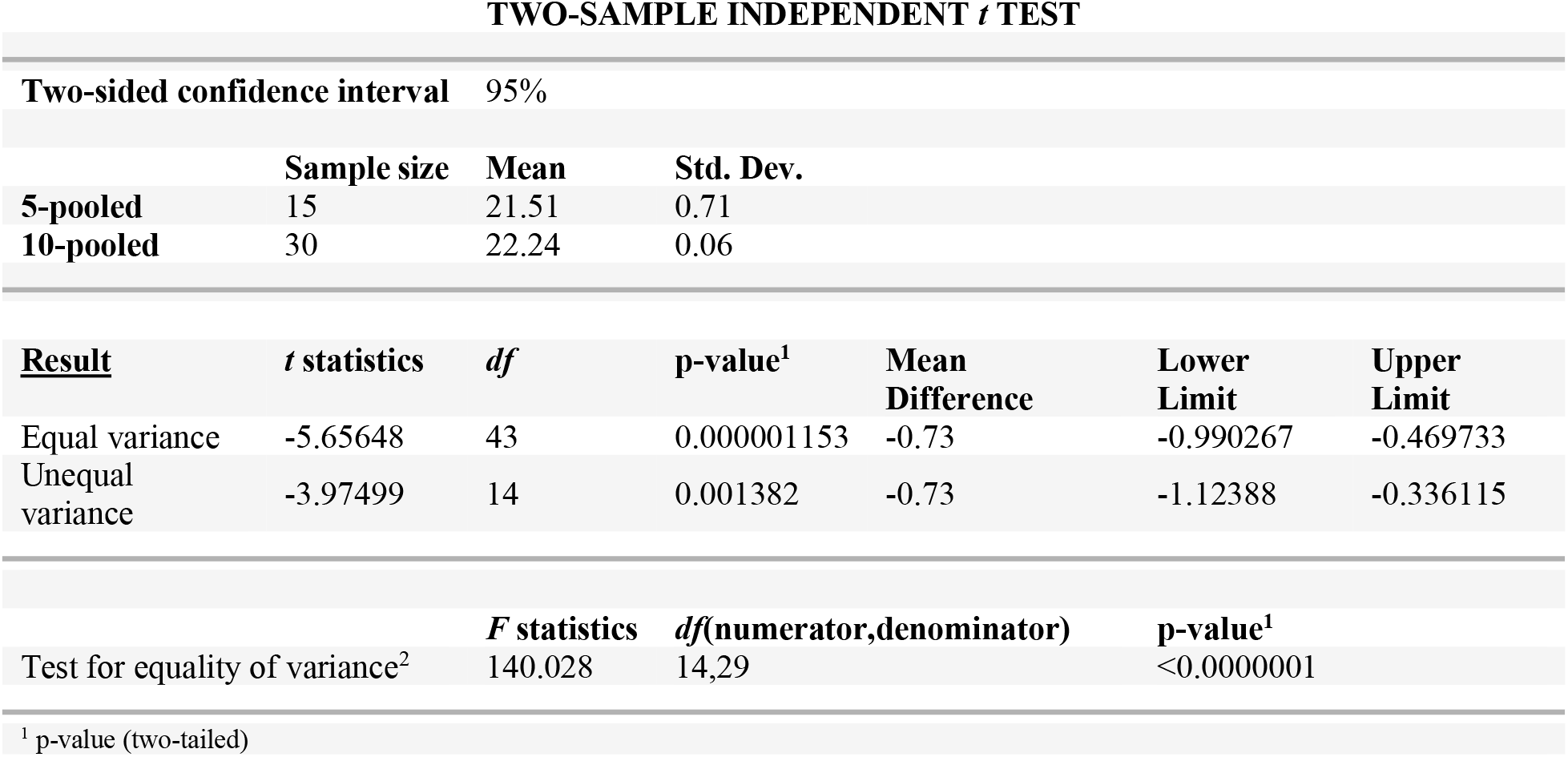
Comparison of mean Ct value of 5- and 10-pooled samples in post extraction phase with a Ct value of <24.

**Supplementary Table S7.** Comparison of mean Ct value of 5- and 10-pooled samples in post extraction phase with a Ct value of 25-28.

| TWO-SAMPLE INDEPENDENT *t* TEST |  |  |  |  |  |  |
| --- | --- | --- | --- | --- | --- | --- |
| Two-sided confidence interval |  | 95% |  |  |  |  |
|  | Sample size | Mean | Std. Dev. |  |  |  |
| 5-pooled | 15 | 28 | 0.06 |  |  |  |
| 10-pooled | 30 | 29.4 | 0.16 |  |  |  |
| Result | *t* statistics | *df* | p-value<sup>1</sup> | Mean Difference | Lower Limit | Upper Limit |
| Equal variance | -32.6047 | 43 | <0.0000001 | -1.4 | -1.48659 | -1.31341 |
| Unequal variance | -42.3401 | 41 | <0.0000001 | -1.4 | -1.46678 | -1.33322 |
|  |  | *F* statistics | *df*(numerator,denominator) | p-value<sup>1</sup> |  |  |
| Test for equality of variance<sup>2</sup> |  | 7.11111 | 29,14 | 0.0003708 |  |  |
| <sup>1</sup> p-value (two-tailed) |  |  |  |  |  |  |

**Supplementary Table S8.** Comparison of mean Ct value of 5- and 10-pooled samples in post extraction phase with a Ct value of 29-32.

| TWO-SAMPLE INDEPENDENT *t* TEST |  |  |  |  |  |  |
| --- | --- | --- | --- | --- | --- | --- |
| Two-sided confidence interval |  | 95% |  |  |  |  |
|  | Sample size | Mean | Std. Dev. |  |  |  |
| 5-pooled | 15 | 34.12 | 2.52 |  |  |  |
| 10-pooled | 30 | 33.38 | 0.88 |  |  |  |
| **Result** | ***t* statistics** | ***df*** | **p-value<sup>1</sup>** | **Mean Difference** | **Lower Limit** | **Upper Limit** |
| Equal variance | 1.4541 | 43 | 0.1532 | 0.74 | -0.28631 | 1.76631 |
| Unequal variance | 1.10414 | 16 | 0.2859 | 0.74 | -0.680758 | 2.16076 |
|  |  | ***F* statistics** | ***df*(numerator,denominator)** | **p-value<sup>1</sup>** |  |  |
| Test for equality of variance<sup>2</sup> |  | 8.20041 | 14,29 | 0.000002174 |  |  |
| <sup>1</sup> p-value (two-tailed) |  |  |  |  |  |  |

**Supplementary Table S9.**
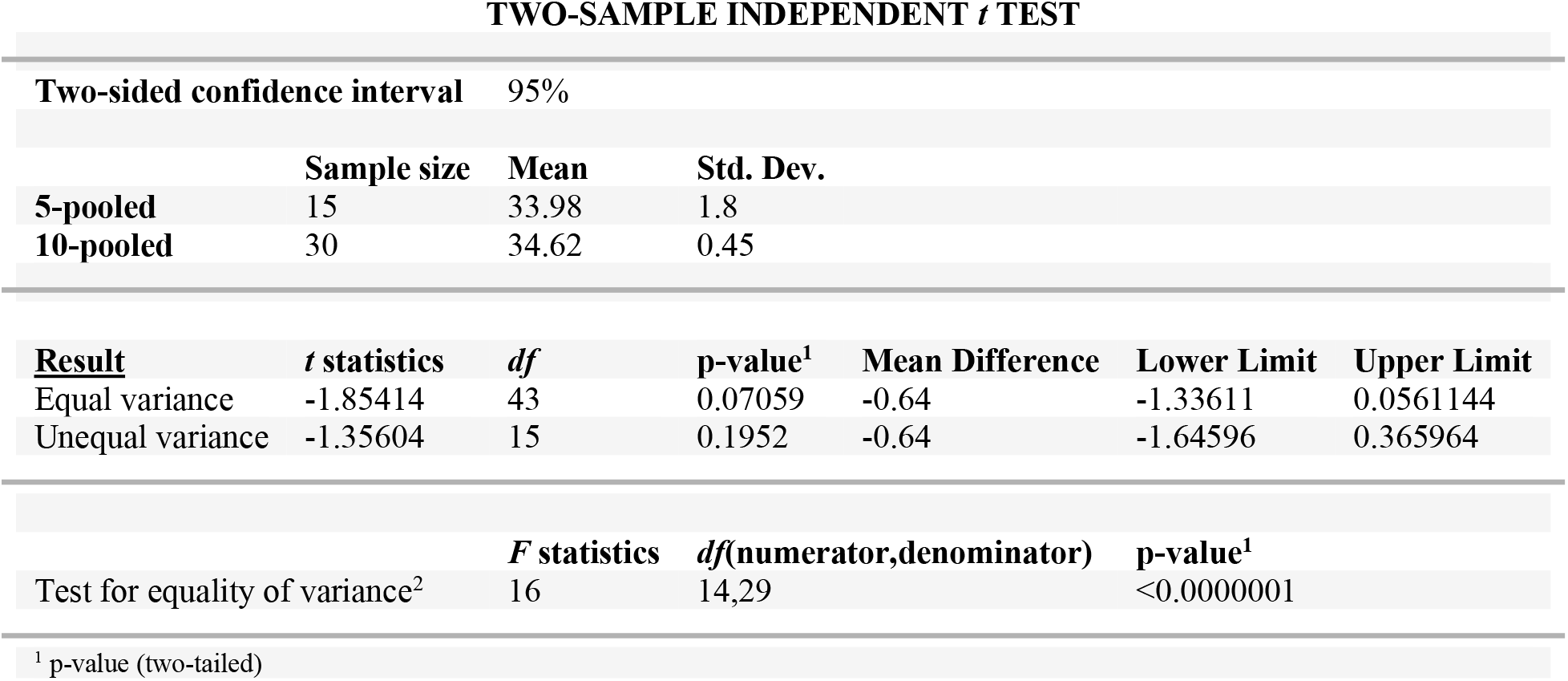
Comparison of mean Ct value of 5- and 10-pooled samples in post extraction phase with a Ct value of 33-36.

**Supplementary Table S10.**
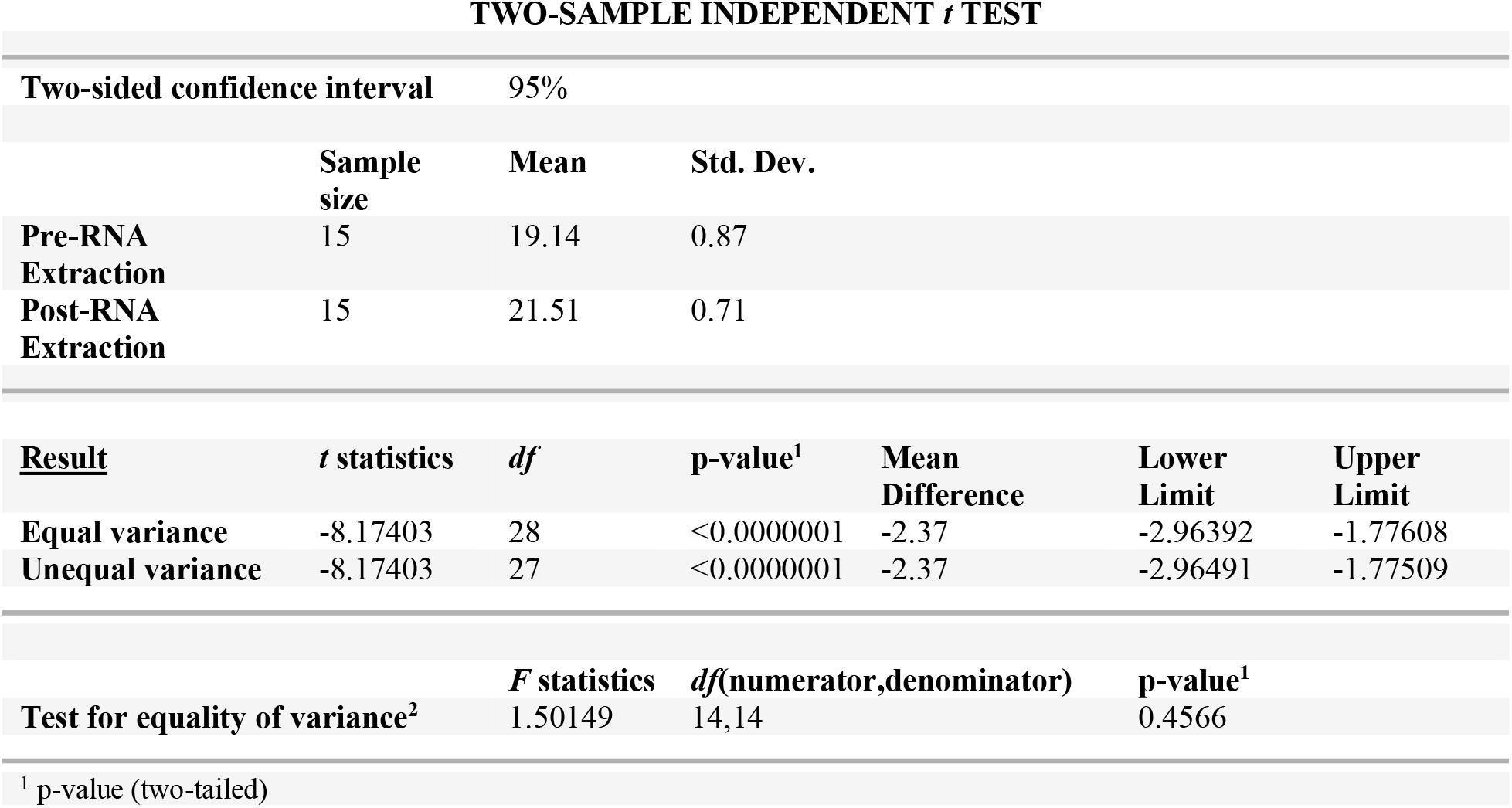
Comparison of mean Ct value of pre- and post-RNA extraction phase in 5-pooled samples with a Ct value of <24.

**Supplementary Table S11.**
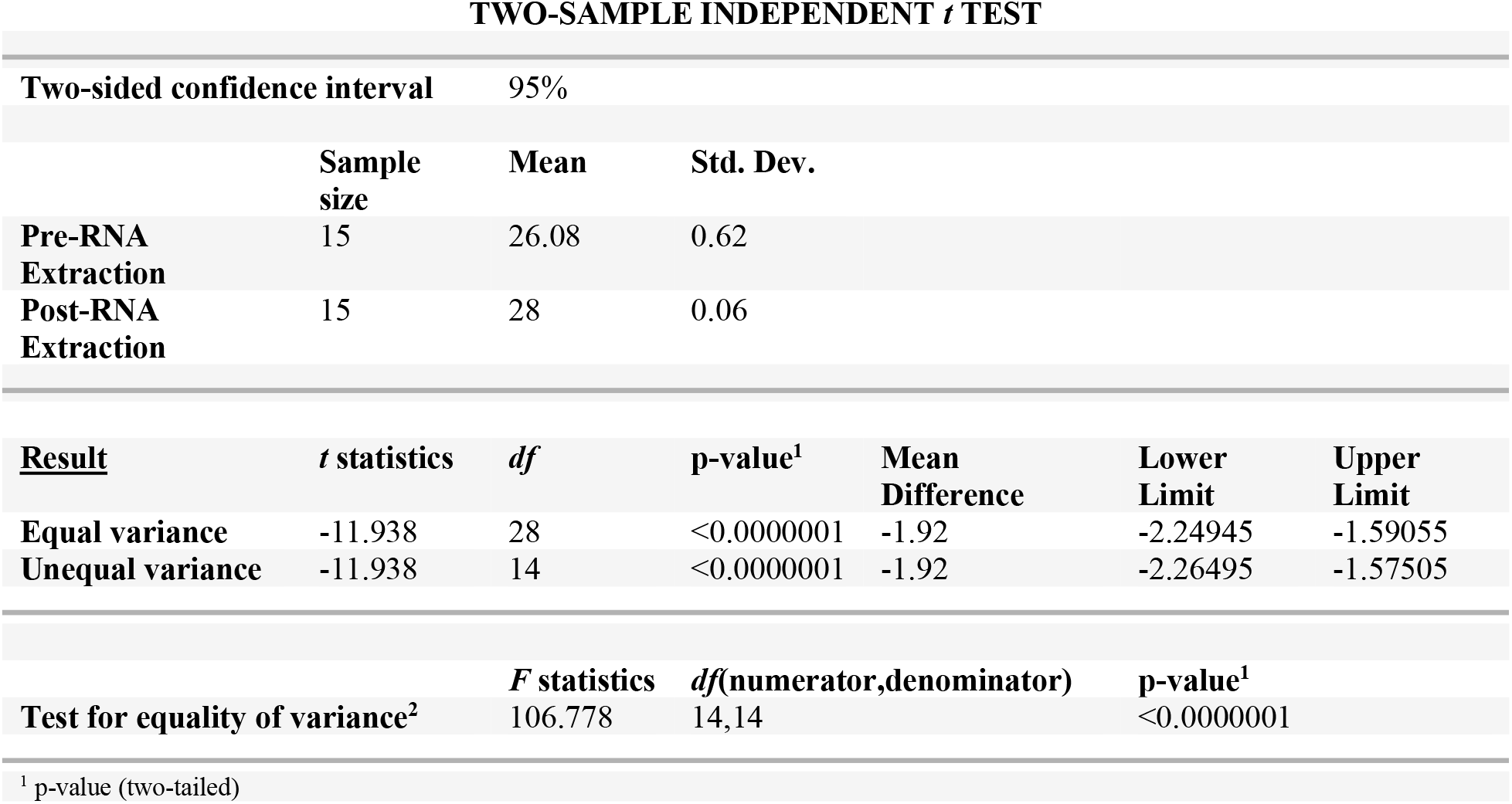
Comparison of mean Ct value of pre- and post-RNA extraction phase in 5-pooled samples with a Ct value of 25-28.

**Supplementary Table S12.**
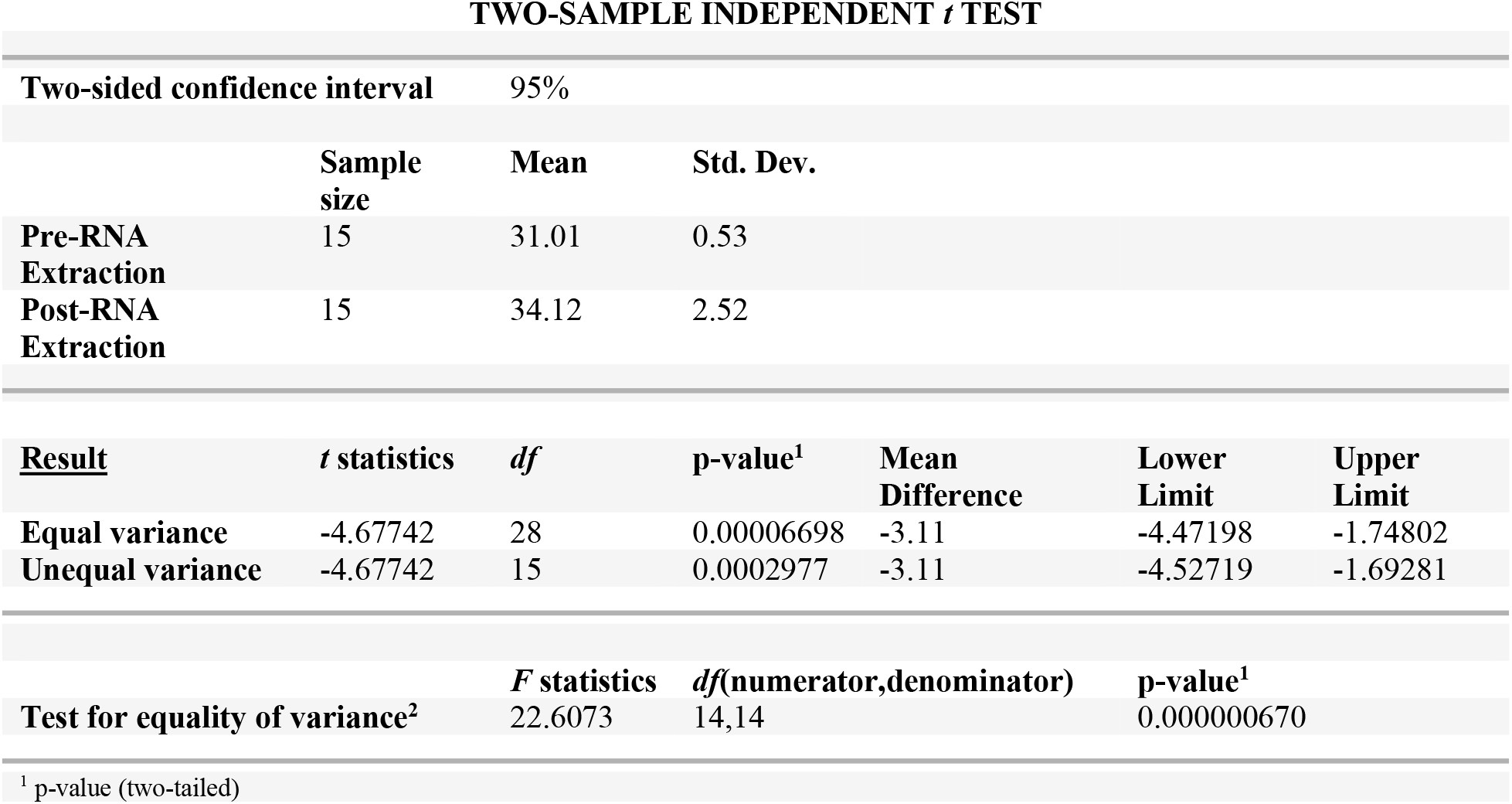
Comparison of mean Ct value of pre- and post-RNA extraction phase in 5-pooled samples with a Ct value of 29-32.

**Supplementary Table S13.**
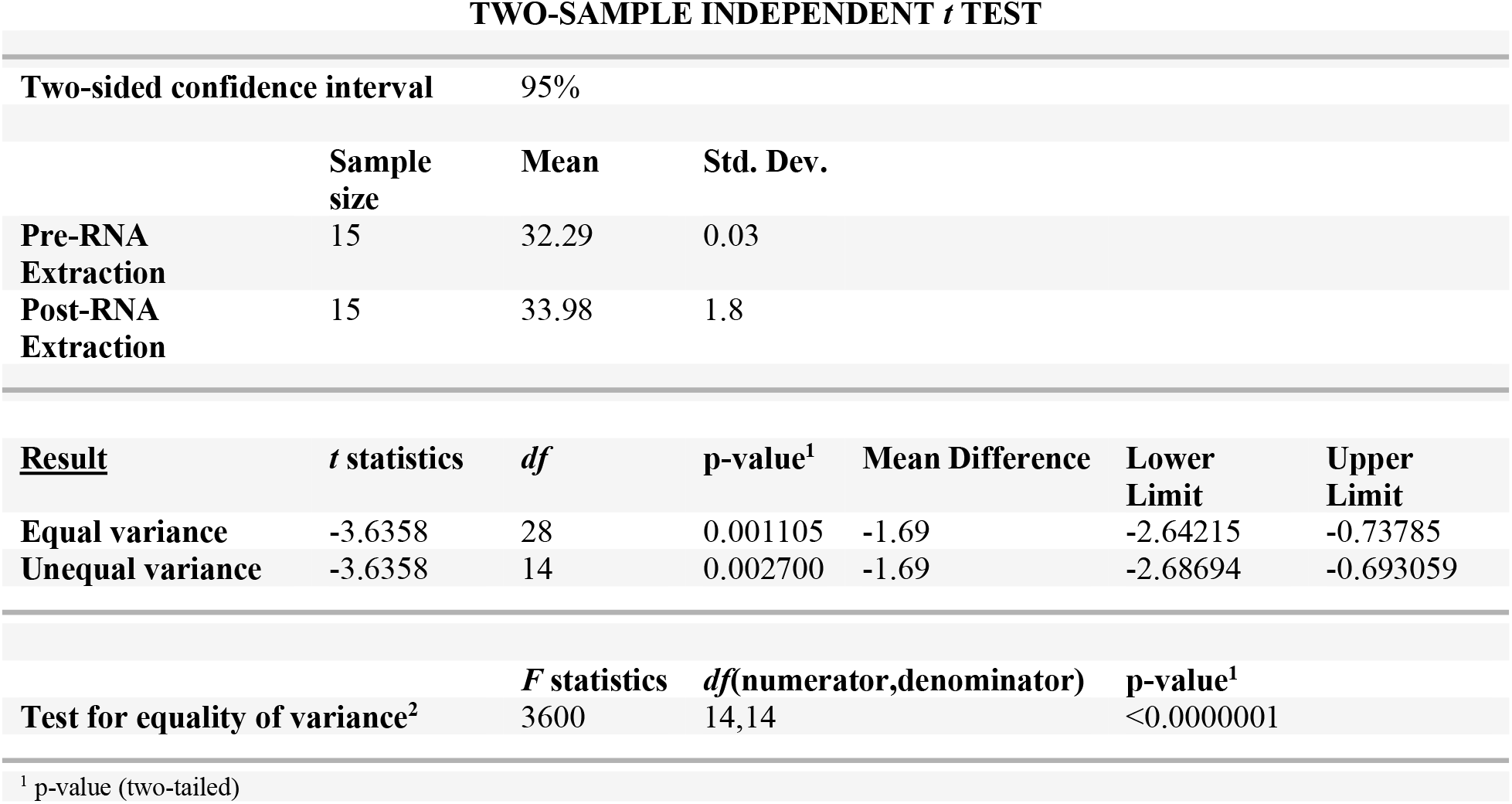
Comparison of mean Ct value of pre- and post-RNA extraction phase in 5-pooled samples with a Ct value of 33-36.

**Supplementary Table S14.**
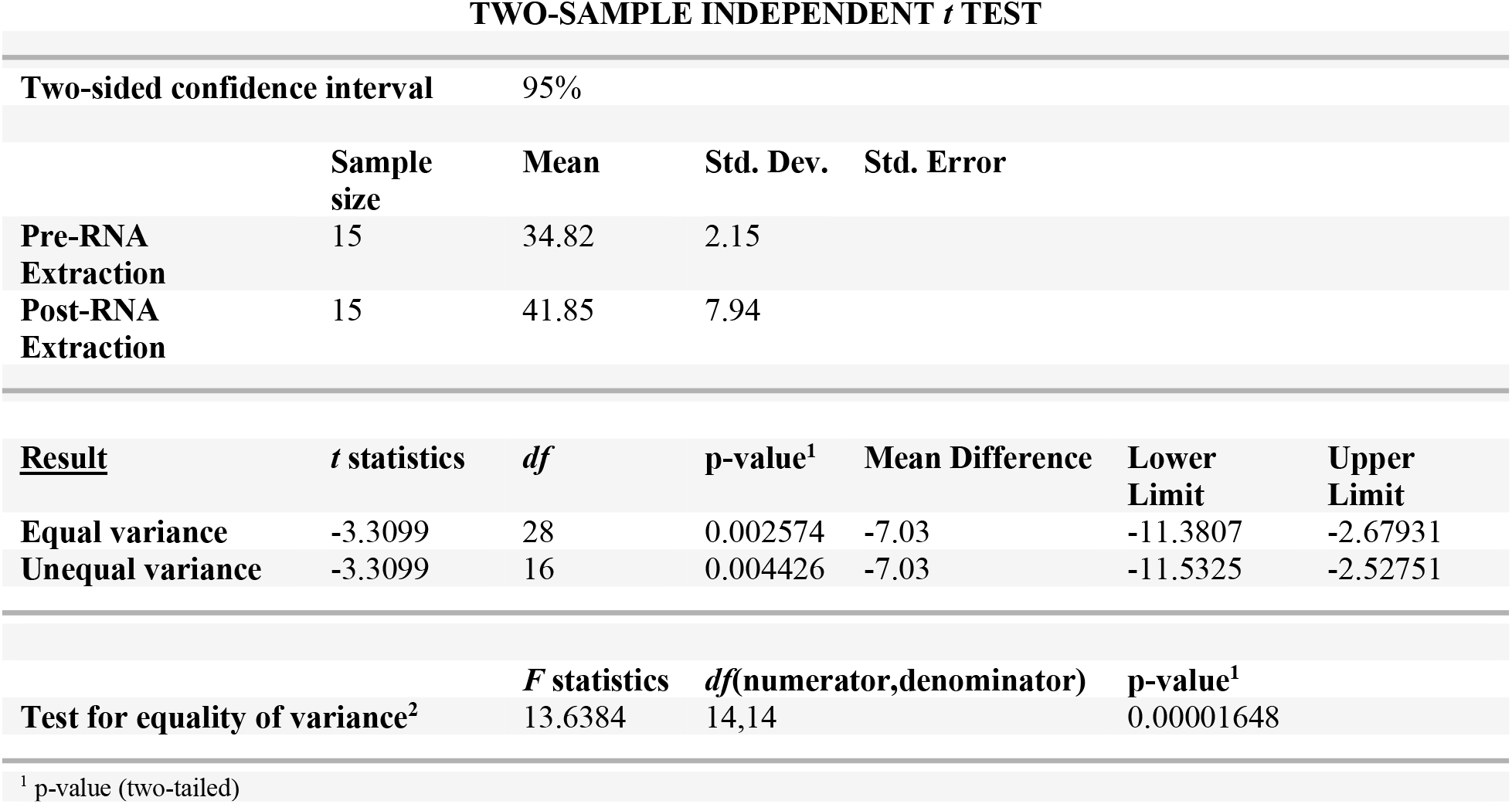
Comparison of mean Ct value of pre- and post-RNA extraction phase in 5-pooled samples with a Ct value of 37-40.

**Supplementary Table S15.**
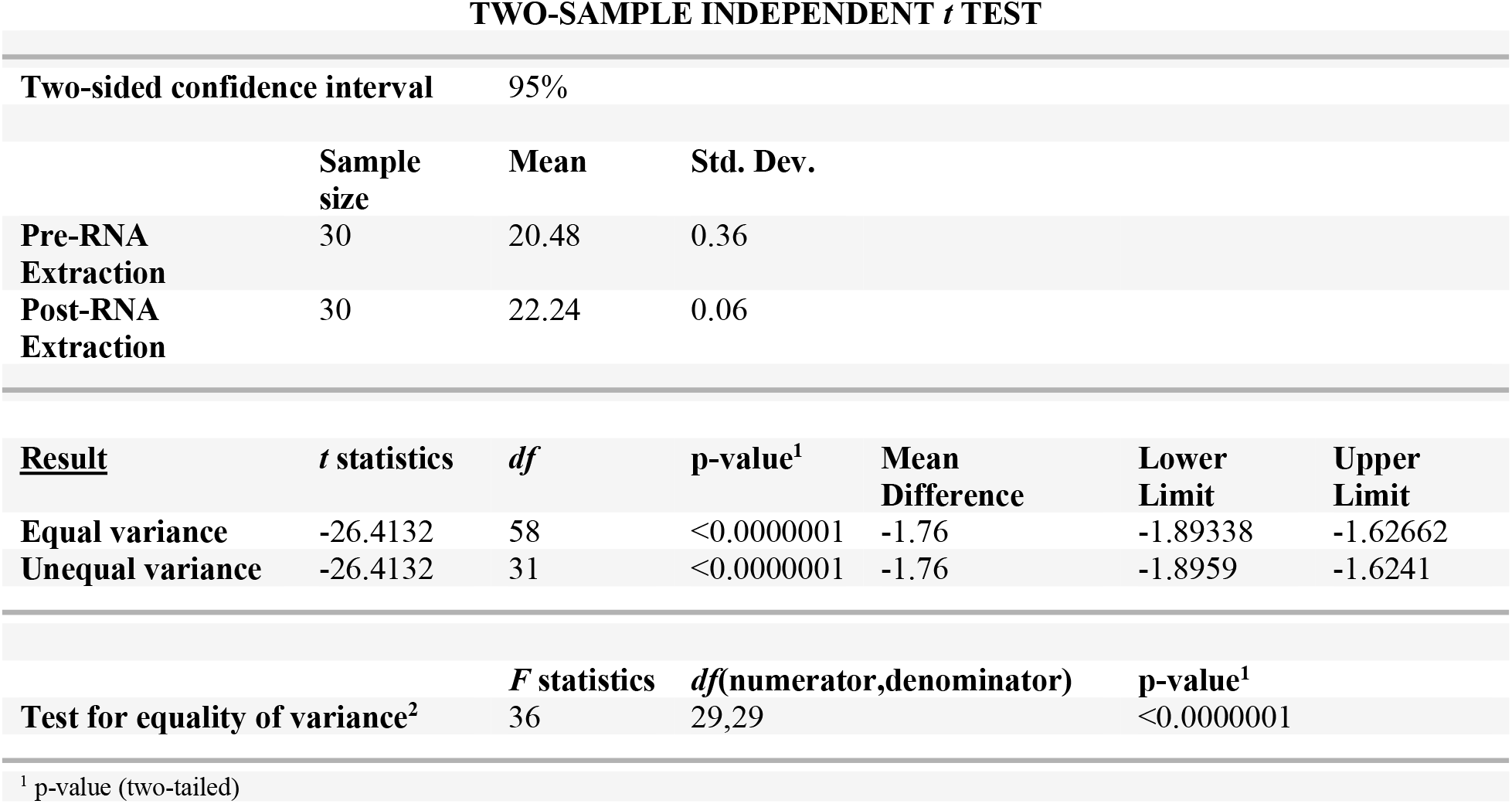
Comparison of mean Ct value of pre- and post-RNA extraction phase in 10-pooled samples with a Ct value of <24.

**Supplementary Table S16.** Comparison of mean Ct value of pre- and post-RNA extraction phase in 10-pooled samples with a Ct value of 25-28.

| TWO-SAMPLE INDEPENDENT *t* TEST |  |  |  |  |  |  |
| --- | --- | --- | --- | --- | --- | --- |
|  | Sample size | Mean | Std. Dev. |  |  |  |
| Pre-RNA Extraction | 30 | 27.16 | 1.15 |  |  |  |
| Post-RNA Extraction | 30 | 29.4 | 0.16 |  |  |  |
| <u>Result</u> | *t* statistics | *df* | p-value<sup>1</sup> | Mean Difference | Lower Limit | Upper Limit |
| Equal variance | -10.5669 | 58 | <0.0000001 | -2.24 | -2.66433 | -1.81567 |
| Unequal variance | -10.5669 | 30 | <0.0000001 | -2.24 | -2.67292 | -1.80708 |
|  |  | *F* statistics | *df*(numerator,denominator) | p-value<sup>1</sup> |  |  |
| Test for equality of variance<sup>2</sup> |  | 51.6602 | 29,29 | <0.0000001 |  |  |

<sup>1</sup> p-value (two-tailed)

**Supplementary Table S17.** Comparison of mean Ct value of pre- and post-RNA extraction phase in 10-pooled samples with a Ct value of 29-32.

| TWO-SAMPLE INDEPENDENT *t* TEST |  |  |  |  |  |  |
| --- | --- | --- | --- | --- | --- | --- |
| Two-sided confidence interval |  | 95% |  |  |  |  |
|  | Sample size | Mean | Std. Dev. |  |  |  |
| Pre-RNA Extraction | 30 | 32.76 | 1.8 |  |  |  |
| Post-RNA Extraction | 30 | 33.38 | 0.88 |  |  |  |
| <u>Result</u> | *t* statistics | *df* | p-value<sup>1</sup> | Mean Difference | Lower Limit | Upper Limit |
| Equal variance | -1.69489 | 58 | 0.09546 | -0.62 | -1.35224 | 0.112236 |
| Unequal variance | -1.69489 | 42 | 0.09750 | -0.62 | -1.35823 | 0.118229 |
|  |  | *F* statistics | *df*(numerator,denominator) | p-value<sup>1</sup> |  |  |
| Test for equality of variance<sup>2</sup> |  | 4.18388 | 29,29 | 0.0002375 |  |  |

<sup>1</sup> p-value (two-tailed)

**Supplementary Table S18.**
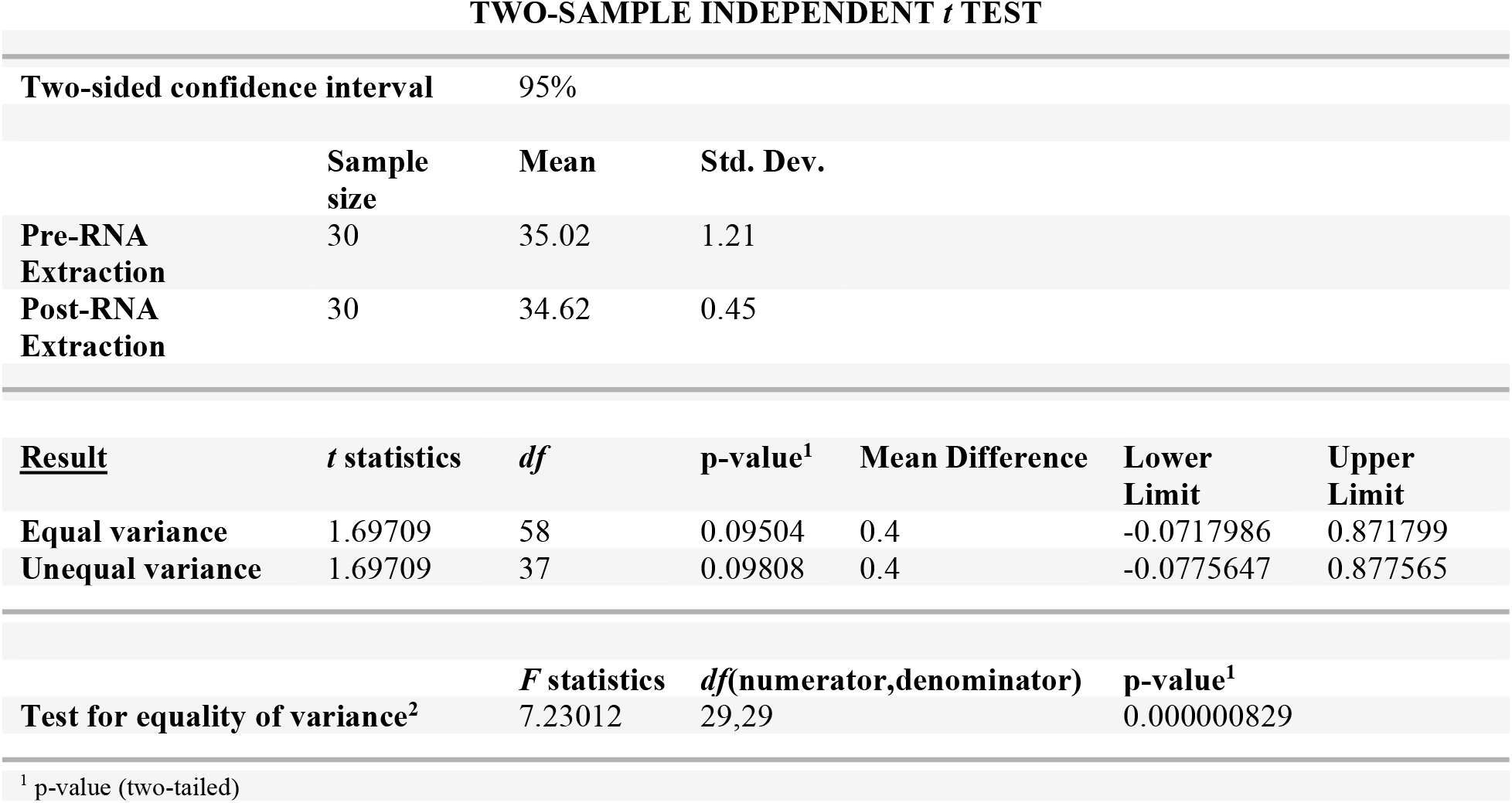
Comparison of mean Ct value of pre- and post-RNA extraction phase in 10-pooled samples with a Ct value of 33-36.

### III. Schematic diagram

**Figure 1.**
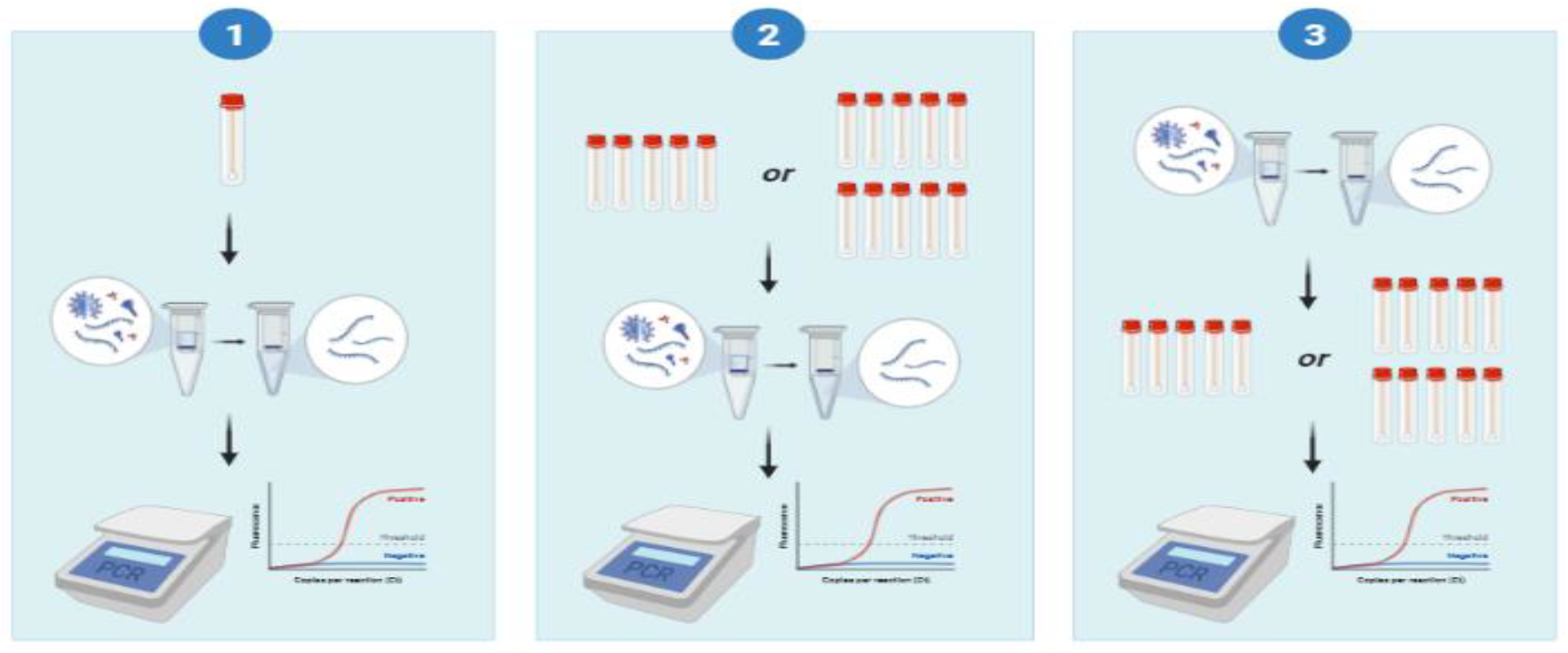
Pooling procedure of known samples. (1) The usual RT-PCR process flow where individual samples underwent RNA extraction prior to RT-PCR testing. This was set as the gold standard of the analysis for this stage. Once Ct values of the individual samples were determined, these were then assigned to represent each of the Ct value ranges given with one positive and four negative samples each pool. (2) Known samples underwent pooling method prior to RNA extraction (5- and 10-pooling) then RT-PCR testing. (3) Known samples underwent pooling method after RNA extraction (5- and 10-pooling) then RT-PCR testing.

**Figure 2.**
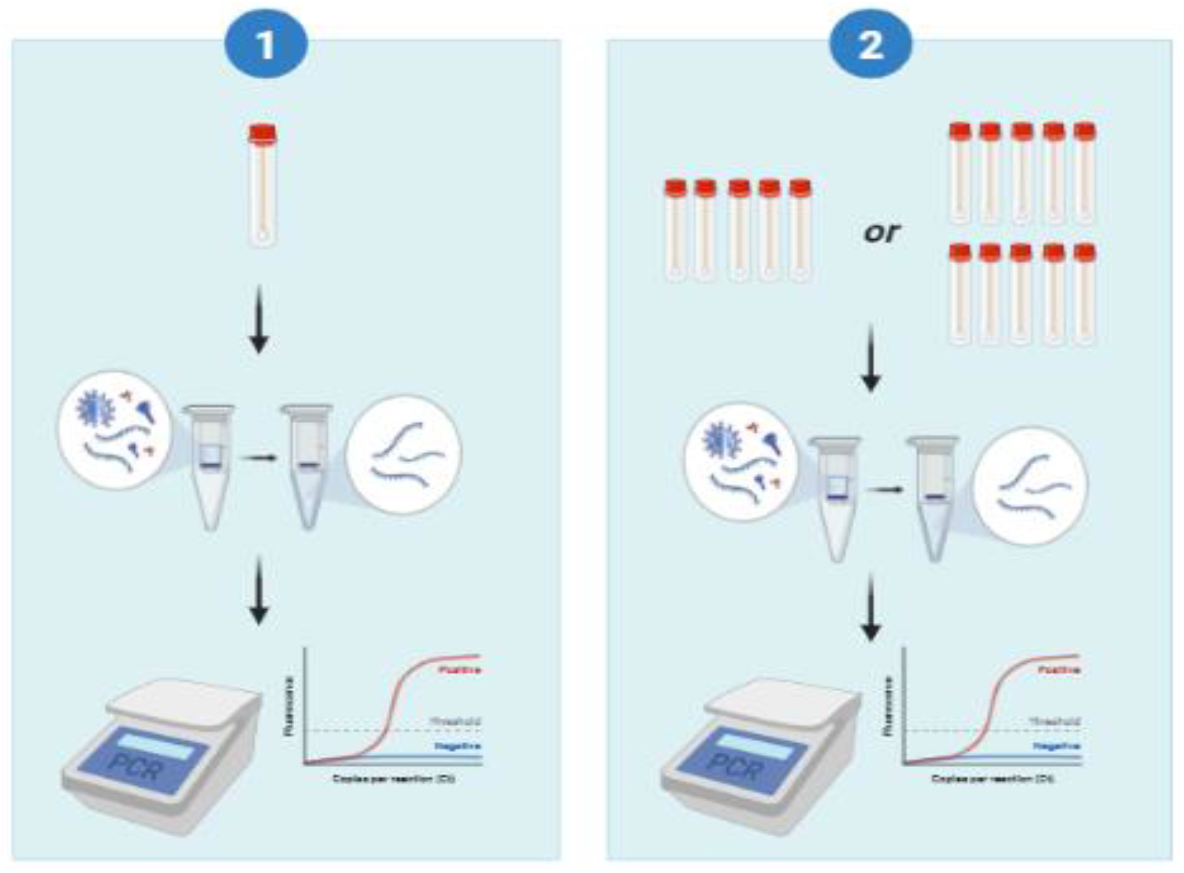
Pooling procedure of unknown samples. This is a schematic diagram of the second stage of the pooled testing method where unknown (clinical) SARS-CoV-2 samples were used. Individual and pooled samples were tested simultaneously. (1) The usual process flow where individual samples underwent RNA extraction prior to RT-PCR testing. This was set as the gold standard for this stage. (2) Validation of the first stage using unknown (clinical samples) pooled prior to RNA extraction.

